# Leveraging tissue-resident memory T cells for non-invasive immune monitoring via microneedle skin patches

**DOI:** 10.1101/2025.03.17.25324099

**Authors:** Sasan Jalili, Ryan R. Hosn, Wei-Che Ko, Khashayar Afshari, Ashok Kumar Dhinakaran, Namit Chaudhary, Laura Maiorino, Nazgol Haddadi, Anusha Nathan, Matthew A. Getz, Gaurav D. Gaiha, Mehdi Rashighi, John E. Harris, Paula T. Hammond, Darrell J. Irvine

## Abstract

Detecting antigen-specific lymphocytes is crucial for immune monitoring in the setting of vaccination, infectious disease, cancer, and autoimmunity. However, their low frequency and dispersed distribution across lymphoid organs, peripheral tissues, and blood pose challenges for reliable detection. To address this issue, we developed a strategy exploiting the functions of tissue-resident memory T cells (T_rm_s) to concentrate target circulating immune cells in the skin and then sample these cells non-invasively using a microneedle (MN) skin patch. T_rm_s were first induced at a selected skin site through initial sensitization with a selected antigen. Subsequently, these T_rm_s were restimulated by intradermal inoculation of a small quantity of the same antigen to trigger the “alarm” and immune recruitment functions of these cells, leading to accumulation of antigen-specific T cells from the circulation over several days. In mouse models of vaccination, we show that application of MN patches coated with an optimized hydrogel layer for cell and fluid sampling to this skin site allowed effective isolation of thousands of live antigen-specific lymphocytes as well as innate immune cells. In a human subject with allergic contact dermatitis, stimulation of T_rm_s with allergen followed by MN patch application allowed the recovery of diverse lymphocyte populations that were absent from untreated skin sites. These results suggest that T_rm_ restimulation coupled with microneedle patch sampling can be used to obtain a window into both local and systemic antigen-specific immune cell populations in a noninvasive manner that could be readily applied to a wide range of disease or vaccination settings.

---

Immune monitoring in both humans and animal models predominantly relies on analyzing peripheral blood. However, the analysis of adaptive immunity in this manner faces a number of limitations. These challenges can be illustrated by considering as one important case the analysis of antigen-specific CD8^+^ T cells, which play crucial protective roles in response to infection, vaccination, and cancer, as well as pathologic roles in autoimmune disease^1,2^: T cells specific for any given peptide antigen can be present at extremely low frequencies in the peripheral blood (e.g., ∼0.05% in vaccinated hepatocellular carcinoma patients^3^ and SARS-CoV-2 mRNA-vaccinated children^4^), even in the presence of antigen stimulation (e.g., growing tumors, chronic infection, or following vaccination)^5,6^. This low frequency combined with practical limitations on blood volumes that can be routinely collected means that biologically-relevant T cell responses often cannot be measured by traditional methods, even using highly sensitive assays such as IFN-γ ELISPOT, and instead are only revealed by culturing T cells in the presence of antigen and expanding the antigen-specific population *ex vivo*^7–10^.

Microneedle (MN) patches are a technology providing a minimally-invasive means to sample immune cells and soluble factors from the skin. These are arrays of tiny sharp projections supported on either a flexible or stiff backing substrate that mechanically penetrate the stratum corneum and epidermis upon application to the skin^11^. MNs have been extensively explored for the delivery of drugs and vaccines into the skin^12–14^. More recently, microneedles have also been exploited to sample interstitial fluid (ISF) from the skin^15–17^. We previously developed a MN patch coated by a thin hydrogel layer, which enabled sampling not only of soluble factors in ISF, but also the recovery of immune cells from the skin^18^. MN sampling is an attractive alternative to skin biopsies or suction blistering, which while used clinically, are invasive and can significantly alter the sampled area^19^. However, these patches only recovered low numbers of cells unless antigen/adjuvant stimuli were included within the sampling hydrogel layer^18^, which complicates translation to human patients. In addition, this sampling strategy primarily recovered tissue-resident memory T cells (T_rm_s). Thus, while very useful for probing tissue-specific immunity, this approach would not necessarily provide a wholistic window on the systemic immune response. We were inspired by the unique properties of T_rm_s to consider whether these skin-resident immune cells could be leveraged to provide enhanced sampling of both the local *and* systemic antigen-specific immune response in individuals.

T_rm_s are one subset of an array of immune cells that are strategically positioned in barrier sites, such as the skin, lungs, and intestines, and act as a localized rapid defense at portals of entry for pathogens^20–22^. In the skin and female reproductive tract, T_rm_ cells actively patrol tissues by extending dendrite-like arms without recirculating through blood or other organs. Upon re-encounter with cognate antigen, T_rm_ can exhibit immediate effector functions and alert the tissue to potential threats, including infections and autoimmune responses^23,24^. One of their most important roles is to “sound the alarm” in response to antigen encounter, via the rapid production of chemokines and cytokines that recruit immune cells from the blood^25,26^. We hypothesized that this alarm function of T_rm_ cells could be exploited to concentrate antigen-specific T cells from the circulation at a selected site in the skin, providing a means to efficiently sample even rare antigen-specific T cells circulating in the peripheral blood using MN patches applied to a site of T_rm_ stimulation. We envisioned T_rm_ could be leveraged in two distinct ways: a first approach would be to intentionally establish a T_rm_ population at a selected site in the skin via intradermal inoculation of small quantities of an antigen/adjuvant in immune animals, followed by subsequent recall of these T_rm_ cells through re-administration of antigen in the skin. This recall step would lead to the recruitment and accumulation of antigen-specific T cells that could be sampled via MN patch application at the stimulation site. A second scenario is to exploit pre-existing T_rm_ established in treatment or disease conditions, such as patients with inflammatory or autoimmune conditions in the skin. In this situation, restimulation of disease-associated T_rm_ through antigen inoculation would similarly recruit circulating immune cells, again facilitating recovery through local MN patch application.

Here we tested these ideas in mouse and human models of vaccination and skin inflammation, respectively, using MN patches carrying a hydrogel cell- and fluid-sampling coating optimized for maximal cell recovery from the skin without the inclusion of antigens and adjuvants within the sampling hydrogel layer itself. In mice, we show mechanistically that induction and recall of T_rm_ cells in the skin enables greatly amplified cell sampling with MN patches, especially antigen-specific CD8^+^ T cells. T_rm_ recall enabled both tissue-resident and circulating cells to be captured from the blood. We demonstrate that hydrogel-coated MN application induces minimal adverse reactions in human volunteers, with the patches being well tolerated for up to 24 hours without any adverse reactions, and present a case study of a human subject with allergic contact dermatitis, suggesting that reactivation of T_rm_ cells in the skin during antigen recall attracts CD4^+^ and CD8^+^ T cells to the skin in humans. MN patches applied to antigen recall sites in this patient showed superior sampling efficiency compared to the established research method of suction blister skin sampling, allowing recovery of T cells, macrophages, natural killer cells, B cells and monocytes without inducing inflammation or aberrant irritation. Microneedle sampling approach may thus enable sampling of rare antigen-specific immune cell populations that would be challenging to detect by conventional means in the setting of vaccination, immunotherapy, or disease monitoring.

## Results

### Hydrogel coatings maximizing cell migration enhance cell sampling by microneedle patches

We previously developed MN patches capable of sampling both ISF and cells from the skin in a minimally invasive manner. These patches are fabricated by melt-molding of polylactide, a biodegradable polymer similar to that used in resorbable sutures^27,28^, to form an array of conical MN projections 550 μm long and 250 μm wide at the base extending from a solid polymer backing. For small-animal studies, we employed patches 2 cm^2^ in area containing 400 MN projections, which are readily applied to the flanks of mice. (**Fig.1a**). These designs were based on MN dimensions shown to be effective for epidermal sampling in humans and rodents^29^. A coating of sucrose and ionically-crosslinked alginate, an FDA Generally-Regarded-as-Safe biocompatible natural polysaccharide that swells strongly in water^30,31^, is cast over the MN projections to serve as the cell/fluid sampling layer. Sampling is carried out by applying the patch to the skin with mild pressure, which causes mechanical penetration of the stratum corneum and entry of MN projections into the epidermis (**Fig. 1b**i). Sucrose in the cell sampling coating (included to augment the mechanical strength of the sampling layer during skin insertion) rapidly dissolves and the alginate layer swells, absorbing ISF and enabling migration of cells into the gel coating (**Fig. 1b**ii). Cells and fluid can then be recovered from the patch by dissolving the alginate layer with EDTA for downstream analysis (**Fig. 1b**iii).

**Figure 1.**
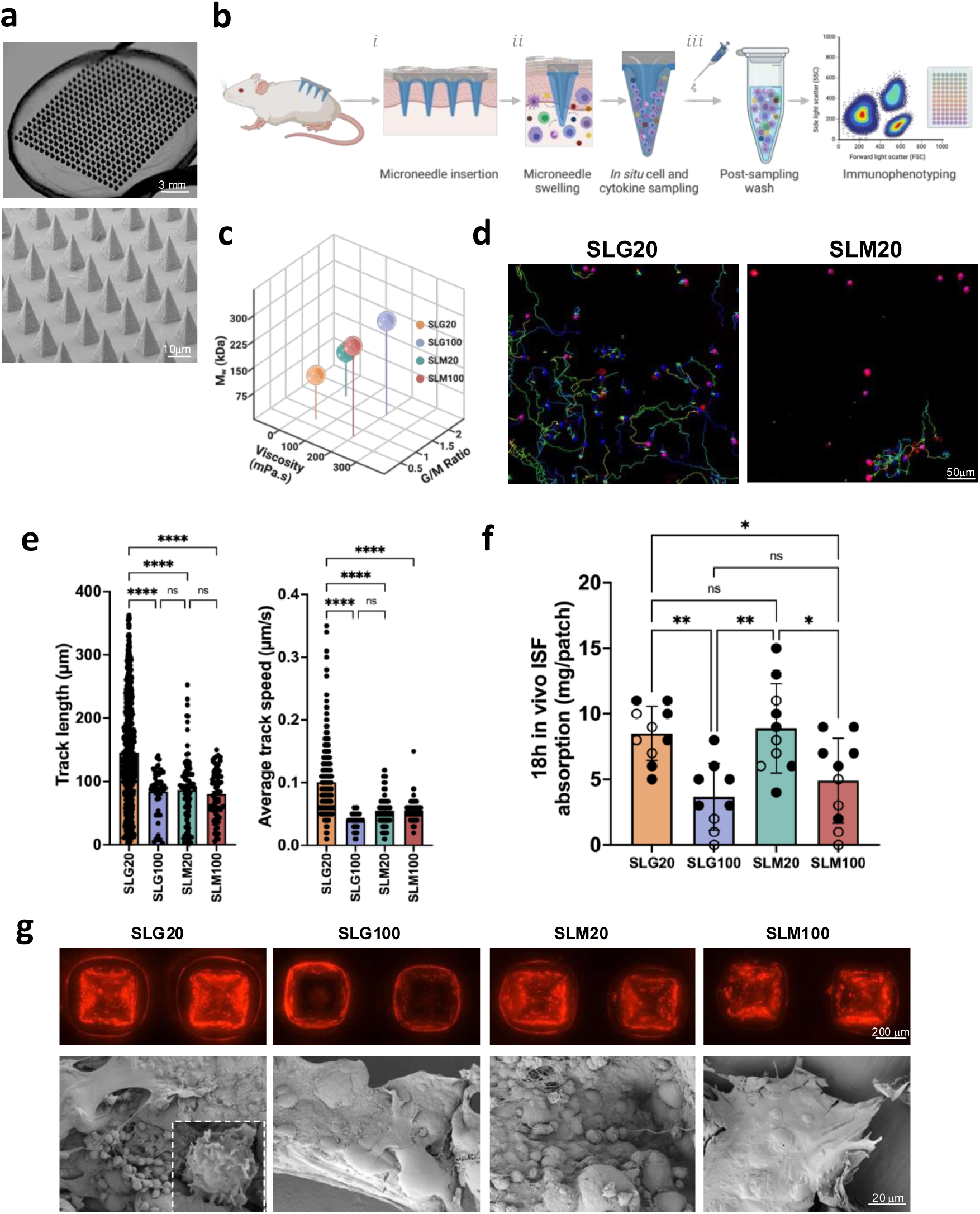
Identifying properties of hydrogel-coated microneedles for optimized cell sampling. **a**, Photographs and scanning electron micrographs of the hydrogel-coated MN patch. **b**, Schematic view of cell and interstitial fluid sampling process with MN array applied to the skin. **c**, Molecular weight, G/M ratio, and viscosity of alginates tested as MN coatings. **d**, Still frames from timelapse microscopy of activated T cells incubated in SLG20 and SLM20 hydrogels *in vitro* showing tracked individual cell paths as colored lines. **e**, Quantification of T cell motility length and average track speed inside different hydrogels. **f**, Comparison of interstitial fluid sampling capacity between microneedles coated with different alginates. **g**, Optical and scanning electron micrographs of the patches after 18 hr of *in vivo* sampling on the skin of OVA-immunized mice and sampled following the scheme of Figure 2b (T_rm_ recall), showing the retention of the alginate layer (labeled with Alexa647 for visualization) and collected cells on the patch. Data shown are means±SEM. ns, nonsignificant; *P<0.05, **P<0.01, ***P<0.001, ****P<0.0001 analyzed by ANOVA, followed by Tukey’s HSD.

Although our previously reported prototype patch design^18^ was functional, we first sought to test whether the composition of the sampling layer could be further optimized to maximize cell recovery from the skin. The sampling layer is formed by dropcasting an alginate and sucrose solution over the MN patch, drying to a solid coating, then crosslinking the alginate with calcium chloride solution followed by a second drying step, to achieve a uniform alginate coating over the MN projections (**Supplementary Fig. S1a-b**). We first tested the influence of the alginate’s molecular weight, guluronic to mannuronic acid ratio (G/M ratio), and viscosity on *in vitro* and *in vivo* fluid absorption, as well as cellular interactions with the hydrogel layer. Four distinct ultra-pure alginates, SLG20, SLG100, SLM20, and SLM100, each with varied physical properties, were tested (**Fig. 1c**). Modifying the G/M ratio and molecular weight of alginate has been demonstrated to alter its physicomechanical properties, including porosity and stiffness^30,32^. *In vitro* bulk swelling of sampling layers prepared with each of these alginates was essentially identical (**Supplementary Fig. S1c**). However, we hypothesized that alginate properties could also affect the ability of cells to migrate within the sampling layer gel, and thereby impact cell collection *in vivo*. To test this, we encapsulated primary mouse T cells in gels prepared from each of the alginates (crosslinked under the same conditions used for sampling layer formation) and tracked cell migration over time by time-lapse microscopy. Strikingly, we observed substantially higher motility and cell migration within SLG20 gels compared to the other alginates, with ∼2-fold greater mean track lengths and ∼2.5-fold greater mean cell speeds than observed for lymphocytes in the other gels (**Fig. 1d, e**).

To test the behavior of these different alginate coatings *in vivo*, we applied patches to the skin of C57Bl/6 mice for 18 hr. In contrast to the *in vitro* swelling measurements, *in vivo* patch sampling revealed distinct quantities of ISF recovered by patches bearing sampling layers prepared with different alginates; sampling layers prepared with SLG20 or SLM20 recovered approximately twice the interstitial fluid of patches prepared with the other two alginates (**Fig. 1f**). Optical and scanning electron microscopy (SEM) imaging revealed distinct differences in the microneedle patches after skin application. Patches coated with SLG100 or SLM100 alginate showed numerous microneedles that had lost their alginate coating following application to skin, resulting in fewer sampled cells accumulating on them (**Fig. 1g**). In contrast, patches coated with SLG20 retained their alginate layer on all microneedle projections and many more cells could be observed associated with the hydrogel coating (**Fig. 1g**). We thus focused further studies on SLG20 as the sampling layer polymer.

We next assessed the impact of the degree of calcium crosslinking, sucrose content, and alginate coating thickness on the behavior of the sampling patches. Sampling layers crosslinked by addition of CaCl_2_ at < 20 mM were too fragile to handle upon swelling, while increasing the CaCl_2_ concentration at the coating crosslinking step led to reduced swelling/fluid uptake, which would reduce ISF recovery (**Supplementary Fig. S1d**). Varying sucrose content and alginate layer thickness, we found that the sucrose content of the sampling layer had only a weak effect on layer swelling/fluid uptake, while thicker alginate layers as expected swelled and absorbed a greater total quantity of fluid *in vitro* (**Supplementary Fig. S1e**). However, MN patches prepared with a thicker vs. thinner sampling layer recovered equivalent numbers of T cells when applied to murine skin (**Supplementary Fig. S1f**), likely due to a limited ability of cells to migrate deep into the sampling layer in the limited time the patches are applied to the skin. Altogether, these studies led us to focus on cell/fluid sampling patches prepared with sampling layers cast from 0.6 wt% SLG20 alginate/2.5 wt% sucrose solutions, approximately 10 µm thick in their final dried state.

### Stimulation of local tissue-resident memory T cells augments skin patch MN cell sampling

In the absence of specific stimuli, MN patches applied to the skin recover few cells and provide little insight into vaccine- or disease-specific immune responses. We hypothesized that the efficiency of sampling antigen-specific lymphocyte populations could be amplified by inducing a population of tissue-resident memory T cells (T_rm_) in the skin, which would serve to actively recruit immune cells from the circulation prior to patch sampling. This concept is schematically outlined in **Fig. 2a**: To establish a T_rm_ population, animals with an antigen-specific immune population of interest (e.g., animals that have been vaccinated with a selected antigen; **Fig. 2a**i) are inoculated intradermally (i.d.) with a small dose of the antigen combined with an adjuvant, to recruit and establish antigen-specific tissue-resident memory T cells at a selected site on the skin (**Fig. 2a**ii). Once established, this antigen-specific T_rm_ population is recalled by i.d. injection of a small dose of the antigen and adjuvant at the same site, which triggers the “alarm” function of T_rm_, leading to rapid production of chemokines that draw a large population of antigen-specific cells to the local skin site^22,33–35^ (**Fig. 2a**iii). MN patch sampling at the site is then used to noninvasively recover and analyze the makeup of the antigen-specific immune response (**Fig. 2a**iV).

**Figure 2.**
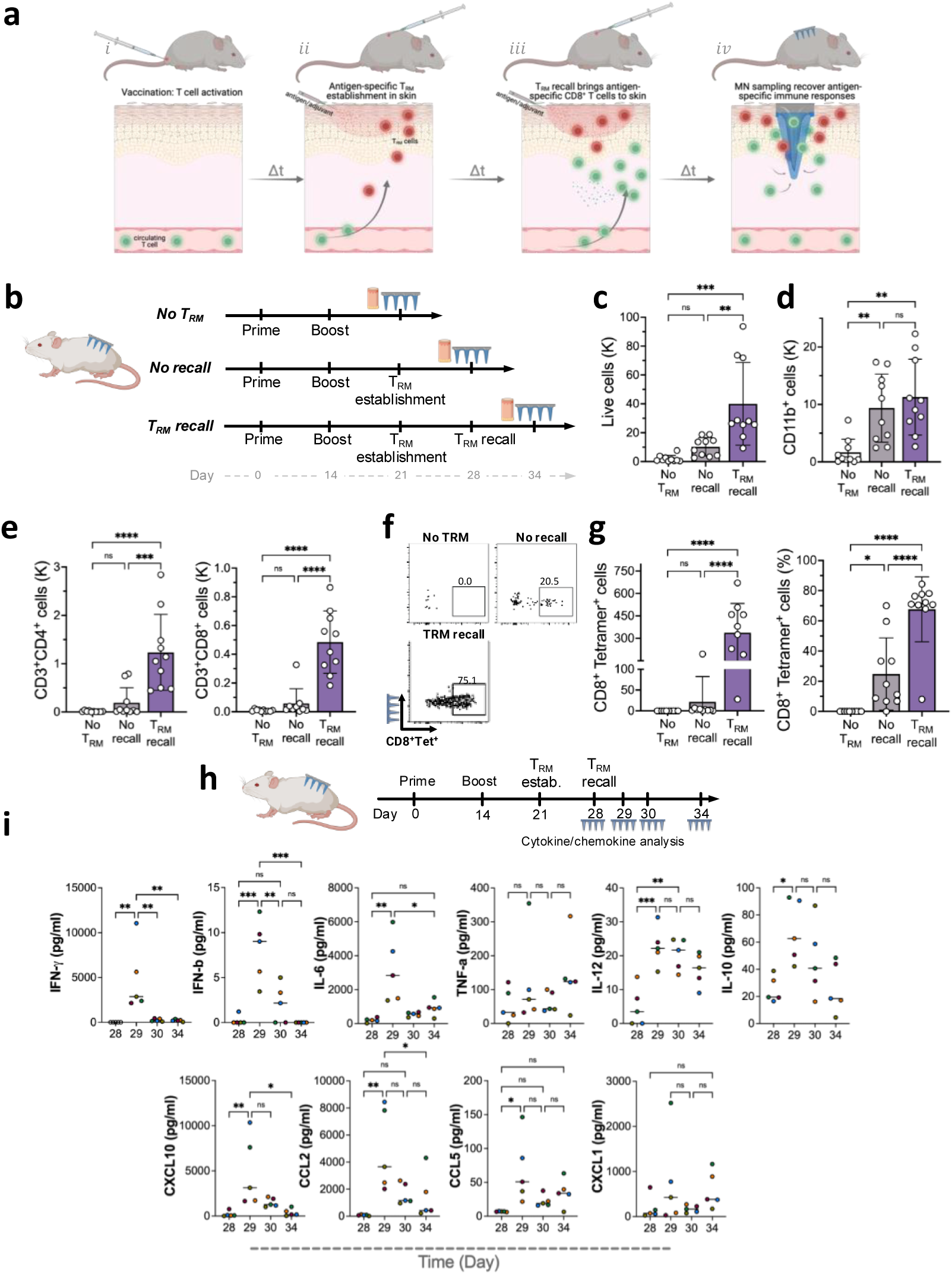
Enhancing microneedle cell sampling via tissue-resident T cell restimulation. **a**, Schematic view of T_rm_ establishment, recall, and recruitment of antigen-specific tissue-resident memory T cells for MN patch sampling at a selected site on the skin. **b**, Study design for establishing T_rm_ populations in the skin and subsequent T_rm_ recall in mice immunized with OVA protein (10 ug per dose) and Lipo-CpG (1.24 nmol per dose). Blood and MN samples were collected 7 days after T_rm_ recall. **c-e**, Enumeration of recovered total live leukocytes, CD3^+^ T cells, T_rm_ cells, and antigen-specific CD8^+^ T cells recovered by MN patches under different sampling regimens. **f,** Representative flow cytometry plots showing SIINFEKL peptide-MHC tetramer staining to identify antigen-specific CD8^+^ T cells collected using MN patches 7 days after T_rm_ recall. **g**, Enumeration of recovered antigen-specific CD8^+^ T cells in the MN patches at T_rm_ establishment and recall steps in comparison with No T_rm_ (*n*=10 animals per group). **h**, study design for longitudinal cytokine sampling post T_rm_ recall step. **i**, Expression of inflammatory cytokines and chemokines induced in the skin measured using multiplexed ELISA analysis of interstitial fluid samples recovered by MN patches following T_rm_ recall. Data shown are means±SEM. ns, nonsignificant; *P<0.05, **P<0.01, ***P<0.001, ****P<0.0001 analyzed by ANOVA, followed by Tukey’s HSD.

We first tested this concept in the setting of immunization against a model antigen and assessed whether T_rm_ stimulation would facilitate enhanced MN sampling of antigen-specific T cells. Groups of mice were primed and boosted with ovalbumin (OVA) and CpG adjuvant, and 7 days post boost, animals were intradermally inoculated with OVA and CpG to establish a T_rm_ population in the skin. One week after T_rm_ establishment, the animals received an intradermal OVA and CpG challenge at the same site to stimulate the local T_rm_, and immune cells recruited to the site under these conditions were assayed by digesting the local skin tissue for analysis using flow cytometry 6 days after the recall dose (“T_rm_ recall” in **Fig. 2b**, **Supplementary Fig. 2a**). We compared immune infiltration at the local site to two other conditions, skin recovered from a site where T_rm_ were induced without the antigen-recall step (**Fig. 2b**, “no recall”), and skin sites without T_rm_ establishment or recall (**Fig. 2b**, “no T_rm_”). Total live cells and CD11b^+^ myeloid cells recovered from the skin in each case showed modest differences, with T_rm_ recall leading to ∼3-fold increases in live cells and myeloid cells recovered from the skin compared to the “no T_rm_” and naïve controls (**Supplementary Fig. 2b, c**). However, T cell infiltration into the analyzed skin sites was dramatically altered by T_rm_ induction: Total CD4^+^ T cells and CD8^+^ T cells recovered from the skin were increased by 12-fold and 5-fold over the “no T_rm_” control, respectively, and OVA-specific CD8^+^ T cells were increased 25-fold over the “no T_rm_” case (**Supplementary Fig. 2d, e**). This enhanced recruitment into the treated skin site depended on T_rm_ restimulation as T cell infiltration was much lower in the “no recall” condition (**Supplementary Fig. 2d, e**), and was likely mediated by recruitment of T cells from the circulation, as only a minority of recovered cells expressed the marker of tissue residency CD103 (**Supplementary Fig. 2f, g**).

Next, we tested cell recovery using MN patches under the same 3 treatment conditions (**Fig. 2b**). In the absence of T_rm_, few live cells were recovered from the MN patches (**Fig. 2c**). When T_rm_ were induced in the skin, a trend toward increased cell recovery at the sampled skin site was observed, but this did not reach statistical significance. However, live cell recovery increased 20-fold over the “no T_rm_” case when the T_rm_ population was recalled with antigen prior to MN sampling (**Fig. 2c**). When we examined the cell types recovered by MN patches, we found that myeloid cell recovery was increased ∼5-fold for both the T_rm_ recall and no-recall groups vs. “no T_rm_” sampling (**Fig. 2d**). By contrast, T_rm_ recall sampling led to 6.5-fold and 8-fold greater CD4^+^ and CD8^+^ T cells recovered, respectively, compared to the no-recall sampling conditions (**Fig. 2e**). This pattern of enhanced T cell recovery by sampling T_rm_-stimulated skin was even more pronounced when we examined the recovery of antigen-specific OVA tetramer^+^ T cells recovered by the MN patches, with T_rm_ recall leading to 16-fold greater numbers of antigen-specific cells recovered compared to the no-recall condition (**Fig. 2f-g**). Notably, the majority of CD8^+^ T cells recovered in the patches using T_rm_ recall were antigen-specific (**Fig. 2g**). Consistent with our initial patch optimization findings, when MN patches with different alginate coatings were employed to sample cells from OVA-immunized mice skin, SLG20 demonstrated superior performance, collecting more live CD45^+^ cells, myeloid cells, CD4^+^ and CD8^+^ T cells as well as antigen-specific OVA tetramer^+^ T_rm_ cells (**Supplementary Fig. 3a**).

Two important parameters in this sampling approach are (i) the interval between T_rm_ recall and patch application and (ii) the duration of patch application. We applied patches in the above sampling experiments 6 days after T_rm_ recall to allow time for robust T cell recruitment to the site from circulation. To confirm that this recruitment period was important, we compared T cell recovery applying patches 1 day or 6 days after T_rm_ recall. Total and antigen-specific T cell recovery was much lower when patches were applied 1 day post T_rm_ recall (**Supplementary Fig. 3b**). When different durations of MN patch application were compared, we observed that the number of live lymphocytes and T cells recovered peaked with an 18 hr duration of patch application to the skin (**Supplementary Fig. 3c**). We thus used SLG20 MN patches, with patches applied for 18 hr at 6 days post T_rm_ recall, for all subsequent cell sampling studies.

To gain insight into the signaling milieu established following T_rm_ recall, we analyzed a panel of inflammatory cytokines and chemokines induced in the skin using multiplexed ELISA analysis of interstitial fluid samples recovered by MN patches applied to treated skin at different time points following T_rm_ recall (**Fig. 2h**). T_rm_ recall induced a coordinated response consisting of induction of an early burst of IFN-γ, IFN-β, and IL-6, at 24 hr which rapidly returned to baseline (**Fig. 2i**). Other chemokines such as CCL5 and CXCL1 showed a trend toward more sustained expression following recall. Thus, “T_rm_ recall” was accompanied by rapid induction of inflammatory cytokine and chemokine induction in the skin and greatly augmented MN sampling of antigen-specific T cells.

Finally, to confirm that the antigen/adjuvant administration used to induce and recall T_rm_s does not itself affect the systemic immune response, we assayed circulating antigen-specific T cell levels following administration of different doses of OVA and adjuvant during the T_rm_ establishment/recall steps. We intentionally administered the T_rm_ recall dose with a 3-week gap after T_rm_ establishment to observe if the intradermal T_rm_ recall injection would lead to a sudden increase in antigen-specific cells in the blood (**Supplementary Fig. S4a**). While we observed that the number of antigen-specific CD8^+^ T cells accumulating in the local site in response T_rm_ recall treatment was dependent on the dose of antigen inoculated in the skin (**Supplementary Fig. S4b-c**) and was amplified only following the final T_rm_ recall inoculation (**Supplementary Fig. S4d**), no significant increase was observed in antigen-specific T cells in the blood following the T_rm_ establishment or T_rm_ recall intradermal injections (**Supplementary Fig. S4e**).

### T cells recovered from T_rm_-stimulated skin reflect the circulating antigen-specific immune response

We hypothesized that T_rm_-recall would lead to recruitment of antigen-specific T cells from the circulation, and thus provide an amplified window into the systemic T cell response. However, stimulated T cells can also proliferate locally^36^, and so it was important to evaluate the role of systemic T cell recruitment vs. local T_rm_ expansion on the makeup of cells recovered by patch sampling. To monitor the migration of antigen-specific T cells to the T_rm_ recall site, we employed photoconvertible KikGR mice^37^ to distinguish sampled cells that were recruited from the blood vs. tissue-resident (**Fig. 3a**). Cells in these mice express the photoreactive KikGR protein in their cytoplasm, which exhibits green fluorescence unless photoactivated by UV light, whereupon a portion of the protein photoconverts to stable red fluorescence (**Fig. 3b**). To employ this tracer for tracking cell recruitment in the skin, OVA-vaccinated mice were inoculated with OVA + CpG adjuvant in the skin to establish T_rm_, and one week later, the sampling site was exposed to UV light to photoconvert skin-resident cells (**Fig. 3a**). Subsequently, T_rm_ were recalled by inoculation of OVA and CpG adjuvant at the same site. Analysis of the skin immediately after photoconversion (0 h) confirmed the near-complete (94%) conversion of these cells from the default green fluorescence of the KikGR protein (“KikGR Green^+^”) to the altered red fluorescent profile (“KikGR Red^+^Green^+^”, **Fig. 3c**). We next vaccinated mice, established T_rm_ by i.d. antigen/adjuvant inoculation (or not), photoconverted the T_rm_ site 1 day before T_rm_ recall on day 28, and sampled the infiltrated cells into the photoconverted area on day 34 via MN patches (**Fig. 3a**). When we analyzed the KikGR reporter expression of recovered cells (**Fig. 3d-e**), we found that in the “no T_rm_” case, the majority of cells sampled– including myeloid cells, CD4^+^ T cells, and CD8^+^ T cells– were KikGR red^+^green^+^ double positive, indicating that they were resident in the skin at the time of photoconversion and that very few recovered cells came from the circulation (**Fig. 3e**). By contrast, the vast majority of cells recovered by T_rm_ recall sampling (including antigen-specific CD8+ T cells) were KikGR green single-positive (**Fig. 3d-e**). This indicates either they were recruited from the circulation or they had proliferated extensively since the photoconversion timepoint.

**Figure 3.**
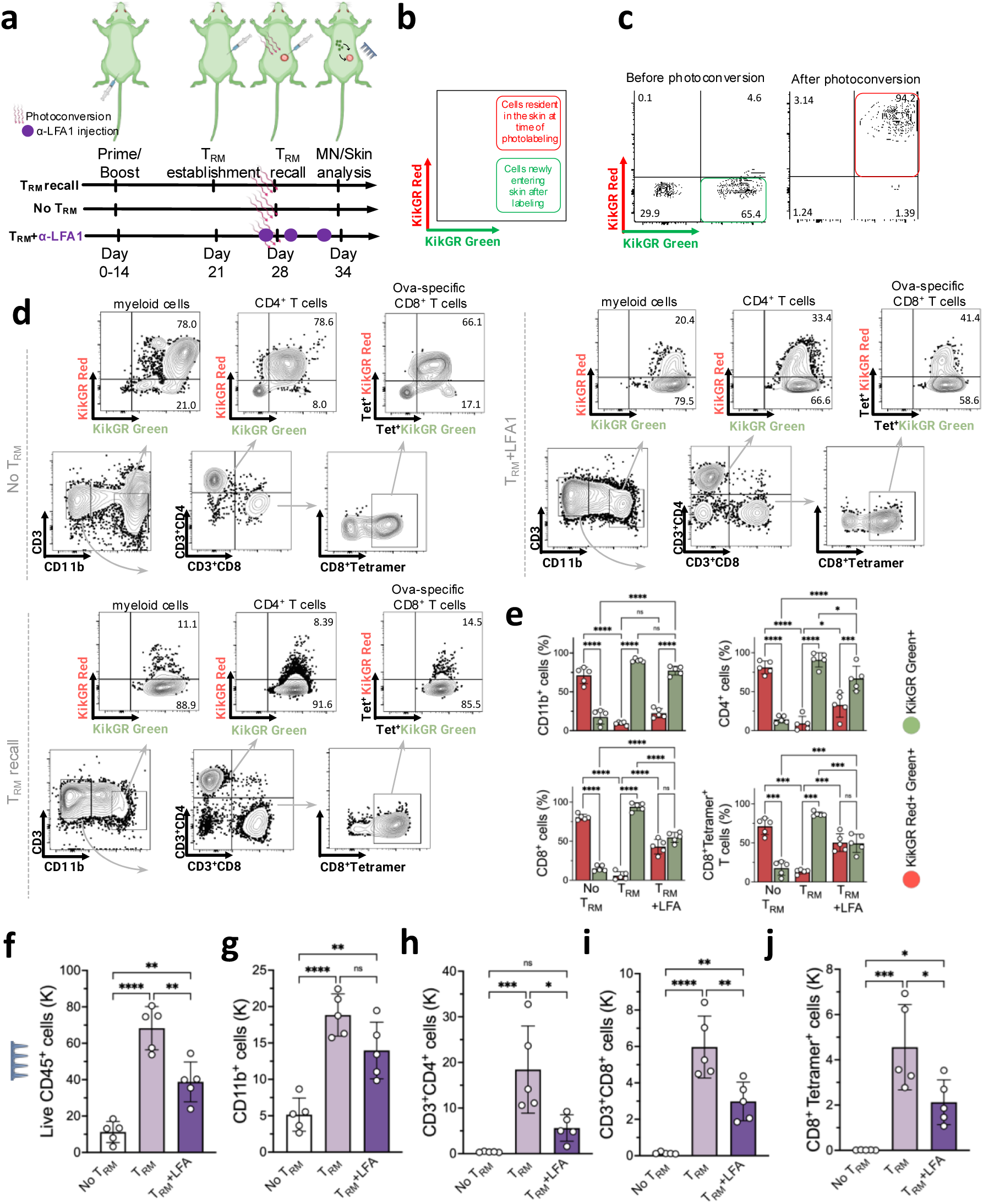
T_rm_ recall leads to recruitment of antigen-specific T cells to the skin from circulation for recovery by MN patches. **a**, Schematic representation of temporal labeling of the skin of the C57BL/6 KikGR mice, photoconverted right before the T_rm_ recall dose (day 28) by violet light exposure on the skin site. **b, c**, Representative flow cytometry plots showing KikGR Red and Green gene expression in skin before and immediately after photoconversion. **d**, Representative flow cytometry plots showing KikGR Red and Green expression by skin infiltrating myeloid cells, CD3, CD4, CD8, and antigen-specific T cells on Day 34. **e,** Quantitation of frequencies of KikGR red^+^green^+^ and green-only^+^ cells by subtype recovered from MN patch sampling following the timeline in 3a. **f-j**, Enumeration of recovered live leukocytes (**f**), myeloid cells (**g**), CD4^+^ T cells (**h**), CD8^+^ T cells (**i**), and antigen-specific CD8^+^ T cells (**j**) 7 days after T_rm_ recall dose. Data shown are means±SEM. ns, nonsignificant, *P<0.05, **P<0.01, ***P<0.001, ****P<0.0001 analyzed by ANOVA, followed by Tukey’s HSD.

To distinguish these possibilities, we carried out sampling under a third condition, where we restimulated T_rm_ in the skin in the presence of systemically administered blocking antibody against Lymphocyte function-associated antigen-1 (LFA-1), as this receptor is important for the homing of T cells from the blood to inflammatory sites^38–40^ (**Fig. 3a**). LFA-1 blockade reduced the total live cells recovered by patch sampling of T_rm_-stimulated skin by ∼2-fold (**Fig. 3f**). While the recovery of myeloid cells showed a slight, non-significant reduction (**Fig. 3g**), LFA-1 blockade significantly reduced CD4^+^ and CD8^+^ T cell recovery by 3-fold and 4-fold, respectively, and decreased OVA-specific CD8^+^ T cell recovery by 2.5-fold, (**Fig. 3h-j**). Further, recovered CD8^+^ T cells and antigen-specific CD8^+^ T cells were ∼50/50 KikGR red^+^green^+^ vs. KikGR green^+^ (**Fig. 3e**). This data suggests that at least ∼50% of the total antigen-specific T cells recovered from T_rm_ recall sampling derived from the circulation, which is a conservative estimate because some proportion of T cell trafficking into inflamed tissues is LFA-1 independent. This data collectively suggests that antigen-specific T cells recovered by T_rm_-recall patch sampling represent both local tissue-resident cells and cells recruited from the circulating blood pool.

### Unveiling T_rm_-driven virus-specific T cell responses with sampling microneedles

While T_rm_ recall was very effective for recovering T cells primed against OVA, this model antigen is highly immunogenic^41^. We thus next sought to test whether T_rm_ recall-based MN sampling could enhance the detection and recovery of antigen-specific T cells primed against a *bona fide* viral antigen. To this end, we synthesized nucleoside-modified mRNA encoding a set of 5 Simian immunodeficiency virus (SIV) T cell epitopes as a model vaccine. This vaccine construct was designed to carry T cell epitopes presented by macaque MHC alleles, but one of the peptides in the mRNA, an epitope termed CL9, can also be presented by mouse class I molecules. A single intramuscular (i.m.) vaccination with lipid nanoparticles carrying this SIV mRNA construct elicited systemic T cell responses recognizing CL9 at a frequency of ∼0.06% among all cells, which could be detected in the spleen by sensitive IFN-γ ELISPOT assays on day 21 (**Fig. 4a**). To evaluate microneedle sampling of this response, T_rm_ cells were established three weeks post mRNA vaccination via i.d. challenge with CL9 peptide and CpG adjuvant, T_rm_s were recalled by i.d. re-administration of CL9+CpG at day 28, and cells were sampled with MN patches at day 34 (**Fig. 4b**, “T_rm_ recall”). We compared T_rm_ recall sampling to a group that only received i.d. peptide/CpG challenge at day 28 post vaccination (**Fig. 4b**, “T_rm_, No recall”). This control condition was intended to establish T_rm_ and provide the same inflammatory stimulus to the skin just prior to MN patch sampling as done in the “T_rm_ recall” group (but without recall stimulation), to account for nonspecific cell recruitment effects of the peptide/adjuvant injection. Skin biopsies at day 34 revealed that the T_rm_ establishment enriched the presence of CD8^+^CD69^+^ and CD8^+^CD103^+^ T_rm_ cells (**Fig. 4c, d**). However, following the T_rm_ recall, we observed an increased number of CD69^-^CD103^-^ cells recruited from the circulation to the site (**Fig. 4d**).

**Figure 4.**
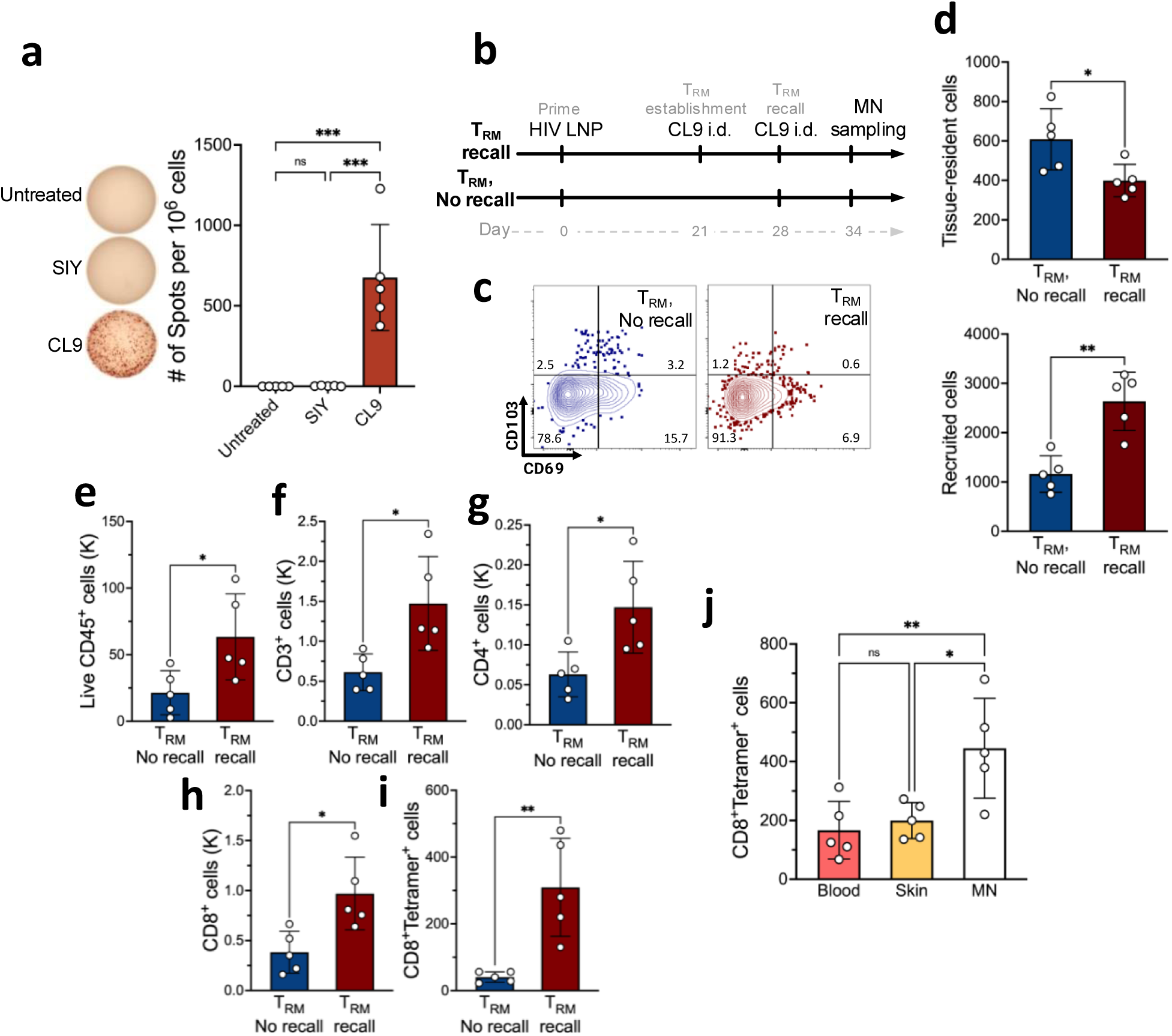
T_rm_-enabled MN patch sampling of virus-specific T cells primed by mRNA vaccination. **a**, ELISPOT analysis of antigen-specific IFN-γ-producing T cells from spleens of mice immunized with mRNA encoding SIV epitopes on day 21 post immunization (*n* = 5 animals/group). **b**, Study timeline comparing MN patch sampling under T_rm_ recall vs. “T_rm_, no recall” conditions. **c**, Representative flow cytometry plots showing expression of tissue residence phenotypic markers for cells recovered from MN patches 7 days after T_rm_ establishment. **d**, Enumeration of tissue-resident (CD103+CD69+, CD103+CD69-, CD103-CD69+) or non-tissue-resident recruited (CD103-CD69-) CD8^+^ T cells recovered from MN patches with or without T_rm_ recall stimulation. **e-i**, Quantitation of total live leukocytes (**e**), total T cells (f), CD4+ T cells (g), CD8+ T cells (h), and antigen-specific CD8+ T cells recovered under recall or no-recall conditions. **j**, Quantitative comparison of antigen-specific CD8^+^ T cells recovered via blood draw (100 ul blood draw), skin biopsy (6 mm punch biopsy), or MN patches. Each data point represents an individual mouse. Data shown are means±SEM. *P<0.05, **P<0.01, analyzed by Student’s t-test or ANOVA, followed by Tukey’s HSD.

MN sampling of animals from these two groups revealed that T_rm_ recall increased the recovery of total live cells, CD3^+^ T cells, CD4^+^ T cells, and CD8^+^ T cells (**Fig. 4e-h**). Most strikingly, CL9 tetramer^+^ CD8^+^ T recovery was increased 7.5-fold in the T_rm_ recall group vs. the T_rm_ no-recall case (**Fig. 4i**). Comparing to a standard mouse blood draw and standard clinical skin punch biopsy (6 mm)^46,47^, patch sampling with T_rm_ recall enabled >2-fold greater recovery of live antigen-specific T cells (**Fig. 4j**). Thus, in response to *bona fide* viral antigen vaccination, T_rm_ recall sampling with MN patches allows for greater recovery of live antigen-specific T cells than traditional blood sampling, and unlike blood sampling, allows recovery of both circulating and tissue-resident T cells.

### Tracking T_rm_ cells and cytokines in human allergic contact dermatitis

The studies above demonstrate a strategy of inducing antigen-specific T_rm_ populations at a selected skin site to enable sensitive sampling of rare circulating T cell populations. A second scenario in which T_rm_ biology can be leveraged for skin MN patch immune monitoring is to use pre-existing disease-induced T_rm_ to locally recruit antigen-specific T cell populations to the skin for sampling. As this approach is compatible with existing clinical practice in dermatologic diseases, we set out to test this concept directly in human volunteers. For human skin sampling, we fabricated square patches with a larger surface area of 4 cm^2^, bearing 400 microneedle projections. The polylactide patch was attached to an adhesive backing to hold it in place following application (**Fig. 5a**). First, under an IRB-approved protocol, we tested microneedle patches for their ease of application and tolerability for an application time from 20 min up to 18 hr for a collection of male and female volunteers covering a broad age range and evaluating a number of different application sites (**Fig. 5a-c**). Clinical records revealed only minimal redness post-patch application, dissipating within an hour, on different skin tones with no instances of bleeding, swelling, or adverse reactions observed (**Fig. 5a**).

**Figure 5.**
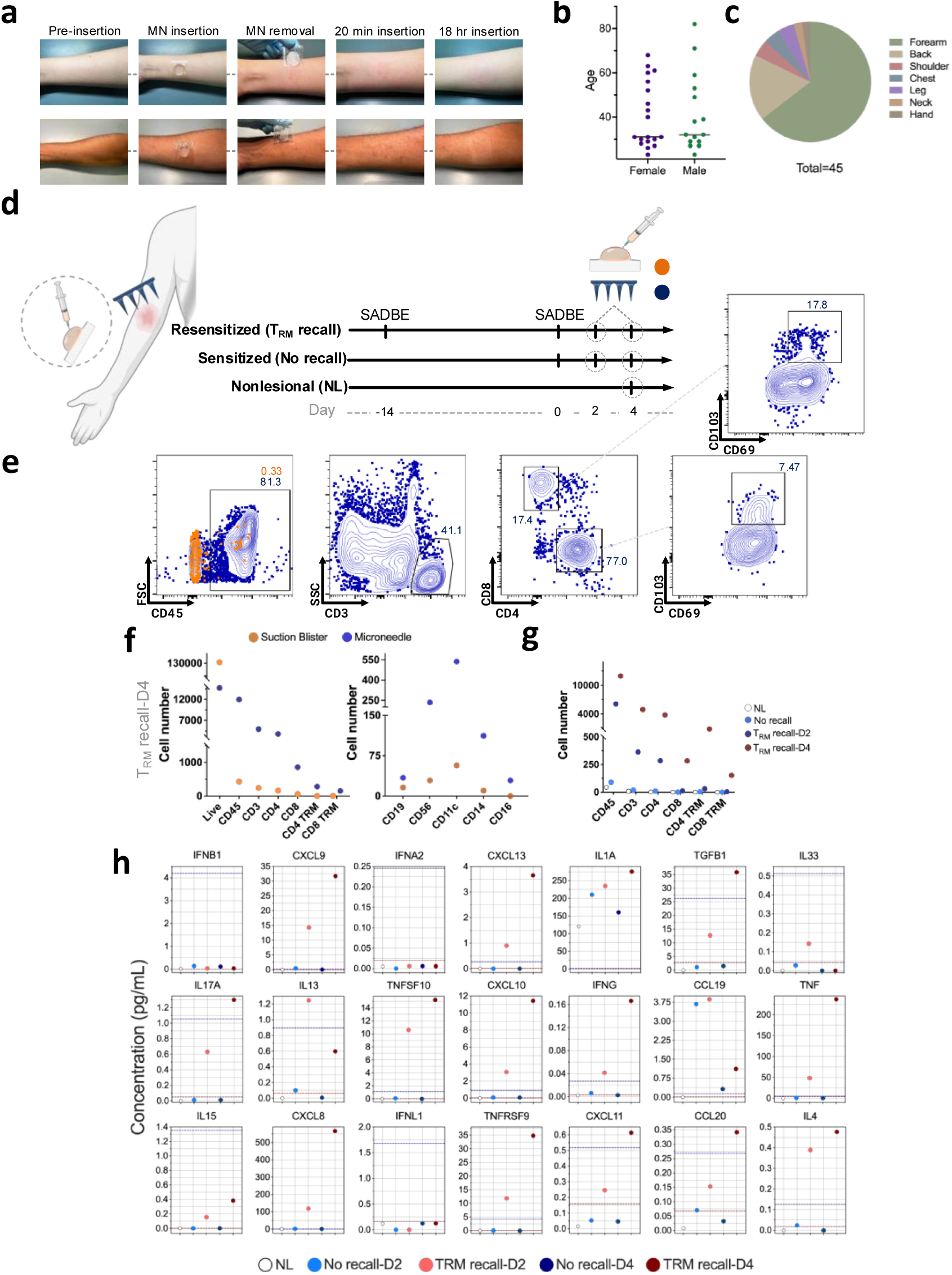
Cell and cytokine sampling in human patient with allergic contact dermatitis. **a**, Representative photographs of pre- and post-MN patch application on the forearm of human volunteers. **b**, **c**. Tolerability of MN sampling was assessed on a cohort of volunteers. Shown is the breakdown of volunteer gender and age (**b**) and skin areas tested (**c**). **d**, Human patient undergo SADBE-induced allergic contact dermatitis with sites previously exposed (14 days before reexposure) to SADBE. Suction blister and microneedle samples were collected 2 and 4 days after SADBE treatment. A non-SADBE exposed site was selected as nonlesional skin. **e**, Representative flow cytometry plots showing expression of immune cell markers in ISF collected from MN patches and suction blisters. **f**, Enumeration of recovered total live, CD45, CD3, CD4, CD8, CD4^+^ T_rm_ and CD8^+^ T cells as recovered by MN patches or suction blister sampling on day 4 of the “T_rm_ recall” condition. **g**, Comparison of cell yields from MN patches applied at 2 vs. 4 days post-T_rm_ recall, alongside nonlesional skin and no-recall control groups. **h**, Olink proteomics data showing temporal changes in skin-associated proteins collected via MN patches.

Tissue-resident memory T cells play an important role in skin allergy, accumulating at skin sites in the initial sensitization phase by an allergen and driving local inflammation on re-exposure to the antigen^48–51^. In a proof-of-concept study, we compared cell recovery obtained from MN patches applied to a human subject after an initial sensitization with the potent contact allergen not found in the natural environment, SADBE^52,53^. This initial exposure represents a “No recall” condition, as there is no chance of prior exposure, and SADBE is safe for such studies due to the minimal risk of accidental re-exposure in the future. We then compared the No-recall case to cell recovery after a T_rm_ recall response, induced by re-exposing the same subject to SADBE 2 weeks later. As an addition control, cell sampling carried out on a distal non-allergen-exposed skin site (nonlesional, **Fig. 5d**). We compared MN patch sampling to cells recovered by suction blister sampling, which we previously used for interstitial fluid sampling from the skin^54^, and collected samples 2 and 4 days after SADBE re-sensitization, to assess the impact of recall kinetics on cell recovery. Recovered cells were analyzed by flow cytometry, aiming to specifically identify recruited CD3^+^ T cells and assess markers of tissue-resident cells (CD69, CD103) as well as B cells and various innate immune cell populations (**Fig. 5e, Supplementary Fig. 5a, b**). While suction blister sampling recovered a larger number of live cells, almost none of these were CD45^+^ immune cells (likely, they are keratinocytes, **Fig. 5f**). By contrast, MN patch sampling recovered almost exclusively immune cells, and this included substantial numbers of CD4^+^ and CD8^+^ T cells, as well as NK cells, dendritic cells, and monocytes (**Fig. 5e-f**). Strikingly, effective recovery of T cell populations from the skin was completely dependent on skin re-sensitization, as very few cells were recovered without recurrent allergen exposure, and cell recovery increased when sampling occurred 4 days post re-sensitization vs. 2 days post re-exposure (**Fig. 5g**).

We additionally conducted advanced Olink proteomics assays on interstitial fluid recovered in the MN patches to track longitudinal immune responses, revealing that T_rm_ recall was associated with an increase in many key cytokines involved in the homing and activation of T cells in the skin compared to the control and non-resensitized skin conditions. These cytokines include CXCL8, CXCL9, CXCL10, CXCL11, CXCL13, IFNγ, IL17a, IL13, IL4, IL15, and TNFα (**Fig. 5h**). Interestingly, proteins detected by microneedle sampling correlated well with those obtained using the traditional suction blister method (**Supplementary Fig. S5c**). While suction blistering is common in skin immune profiling in research settings, it has notable limitations, including elevating local skin temperature, inducing post-inflammatory hyperpigmentation, and impracticality for certain demographics such as those with compromised skin integrity. Many subjects also find suction blistering too painful to tolerate, and the procedure typically requires 50-60 minutes, necessitating a separate research visit in a room equipped with the device, which limits its feasibility in standard clinical settings. Furthermore, the negative pressure and elevated temperature used in suction blistering can cause keratinocyte dissociation, potentially affecting the accuracy of results by introducing high numbers of non-immune cells. These factors make suction blistering challenging for remote or frequent longitudinal sampling, highlighting the need for less invasive and more patient-friendly alternatives like MN patches^55–57^. MN patch sampling not only overcomes these limitations, but also enables live cell recovery, enabling deeper insights into immune reactions occurring in the skin.

## Discussion

Here we have combined microneedle patch technology with the biology of tissue-resident memory T cells to enable efficient sampling of both interstitial fluid and live immune cells from the skin from both mouse models and humans. This approach enables the collection of both tissue-resident and circulating antigen-specific T cell populations, and should enable longitudinal sampling of immune responses if desired over time. Our findings demonstrate that leveraging T_rm_ cells can dramatically amplify the recruitment of antigen-specific T cells from the circulation to sites where T_rm_ cells are established, increasing the efficiency of cell recovery over traditional blood sampling. The ability of T_rm_ to orchestrate immune responses that span innate and adaptive immunity is one of many examples by which the immune system shares information between cell types. Comparison with invasive or complex sampling methods, such as skin biopsy and suction blisters, revealed similar or superior results with microneedle patches in both preclinical animal models and in a human case study.

MN patches have previously been utilized for sampling ISF from the skin, primarily for protein biomarker quantification. Some innovative designs incorporate paper reservoirs^58^ or integrate biosensors to perform *in situ* fluoroimmunoassay on-patch^59^, enhancing their analytical capabilities. However, many existing MN designs incorporate non-FDA approved biological components, which hinder their clinical translation. Additionally, some methods rely on external vacuum devices that lack precise control over sampling depth and prevent remote or at-home sampling^60^. This reliance can lead to complications similar to those seen with suction blistering, including the risk of blood contamination in ISF samples^17^. Furthermore, most of these technologies were developed solely for sampling soluble factors in ISF. While there have been reports of cell sampling using hydrogel MNs, such as those based on hyaluronic acid, the yield of recovered cells remains limited^61^. Addressing these challenges, including improving cell recovery efficiency and developing FDA-compliant materials, is crucial for advancing MN sampling technology and enhancing its applicability in clinical settings.

To optimize cell recovery using sampling microneedle patches, we conducted a screening of various hydrogel coatings with diverse physicochemical properties. Our investigation revealed that a softer alginate coating (SLG20) with a lower G/M ratio and molecular weight, factors previously demonstrated to affect the physical, chemical, and biological properties of alginate biomaterials^30,32,62,63^, yielded the highest cell recovery in vivo. The lower G/M ratio in SLG20 contributes to increased swelling capacity, decreased network size, reduced stiffness, and altered diffusion properties. Specifically, alginates with lower G/M ratios exhibit greater swelling due to a higher proportion of the more flexible M groups, which may facilitate better cell capture and release. The decreased network size resulting from shorter junction zones could allow for easier cell penetration and recovery^64^. The reduced stiffness, attributed to the inherent flexibility of M blocks compared to rigid G blocks, may create a matrix more conducive to cell migration and retrieval. Additionally, the lower molecular weight of SLG20 likely contributes to a less entangled polymer network, further facilitating cell movement and recovery^30,65^. These combined characteristics of lower G/M ratio and reduced molecular weight result in a hydrogel with properties that appear optimal for cell sampling and recovery in our microneedle patch system, highlighting the importance of fine-tuning hydrogel properties for specific biomedical applications. This superior performance of SLG20 was attributed in part to the improved retention of this coating on the patch and its ability to promote increased migration and motility of lymphocytes. Previous studies have highlighted enhanced cell aggregation, cell-cell contact, and cell spreading in softer alginate hydrogels^66^. Additionally, the lower positive charge on the surface of low-G alginate^67^ likely facilitates stronger electrostatic binding of SLG20 to the O_2_-plasma-induced negatively charged patch surface, thereby further enhancing cell yield.

In order to sample immune cell populations of interest, we previously mimicked the classical Mantoux test, where vaccinated animals were inoculated intradermally with small amounts of antigen (with or without adjuvant) to recruit antigen-specific T cells into the skin followed by sampling the injected skin site with microneedles^18^. However, this type of local restimulation in the absence of pre-existing T_rm_ relies on local antigen presentation to memory T cells recruited into the skin from the blood in response to the local tissue inflammation, which in turn produce inflammatory cytokines and chemokines over a period of days to a week that recruits additional antigen-specific lymphocytes from the circulation to the tissue site^68^. As shown here, this approach is much less efficient at accumulating cell populations of interest into the skin compared to restimulation of locally-established tissue-resident T cells. To date, T_rm_ cells in peripheral tissues have been assigned multiple functions. For instance, establishment of tissue-resident virus-specific T cells in the vaginal mucosa has been shown to provide enhanced immunity against genital HSV-2 infection^69^. In the female mouse reproductive tract, these memory CD8^+^ T cells, upon encountering cognate antigen *in situ*, trigger a robust local expression of inflammatory chemokines, facilitating the rapid recruitment of unstimulated circulating memory T cells^33^. Further, CD8^+^ T_rm_ cells demonstrate the ability to induce increases in vascular permeability shortly after reactivation, resulting in the extravasation of circulating antibodies into local tissues and increased neutralization of virus ex vivo^70^. Both of these functions of T_rm_ could potentially play a role in the MN sampling process established here.

In a proof-of-concept study focusing on allergic dermatitis, where a patient’s skin was resensitized with a potent allergen, we observed an increase in the abundance of T cells and T_rm_ cells in resensitized patients compared to those exposed to the allergen once and nonlesional skin sites. This data reaffirms the role of T_rm_ cells in skin allergy, contributing to the recurrence of lesions by potentially recruiting CD4 and CD8 T cells to the skin ^48,72,73^. Proteomics analysis of the interstitial fluid recovered from MN patches in this experiment revealed a longitudinal increase in cytokines responsible for homing and activation of T cells, including IL17a, IFNγ, CXCL8, CXCL10, CXCL11, and CCL20^74–76^. While this case study serves as a proof of concept, we did not specifically characterize antigen-specific T cells appearing in the patient’s skin in response to T_rm_ recall; further studies in larger patient cohorts are warranted. Nevertheless, our findings demonstrate that sampling MN patches can facilitate longitudinal, noninvasive monitoring of humoral and cellular responses in human patients, comparable to the suction blister method but without its potential limitations. Blistering, for instance, can elevate skin temperatures, potentially affecting cytokines/chemokines, and may lead to lasting hyperpigmentation. Ethical concerns also restrict the use of blistering approaches, particularly in vulnerable populations such as infants or older adults.

To our knowledge, this study presents the first report of human cell sampling from patients using MN patches. We acknowledge, however, the need for larger, more controlled cohort studies to elucidate the role of T_rm_ in skin T cell recruitment. While our current work focused on enhancing patch sampling capacity and investigating T_rm_ recruitment through conventional immunophenotyping and proteomics, future research will employ advanced techniques like single-cell RNA sequencing to explore underlying mechanisms and T cell repertoires in greater depth. Beyond vaccine settings and skin allergic reactions, we envision broader applications for this technology in monitoring immune responses to infectious agents, predicting disease flares and therapeutic responses in autoimmune conditions, and assessing tissue status in transplantation. Our microneedle patches could be used to sample the injection site reactions, which are often observed in clinical trials of new vaccines, including recent HIV trials. Notably, some of these injection site reactions have been reported in individuals who previously received SARS-CoV-2 mRNA vaccines, although the exact components responsible for these responses—whether lipids, mRNA, PEG, or anti-glycan reactions—remain unclear^71^. Given these considerations, microneedle patches could serve as a non-invasive method to sample and analyze local immune responses in such cases. By targeting injection sites, these patches could facilitate better understanding of the underlying immunological events in vaccinated individuals and could be applied to track immune responses in various vaccine and therapeutic settings without the need for invasive sampling. Moreover, these microneedle patches could be adapted for use in other mucosal sites and cutaneous tumors, offering a practical alternative to current invasive techniques that require in-person visits by medical experts. The high-throughput capability of microneedles facilitates remote monitoring of disease activity for large-scale studies, with particular benefits for sensitive populations such as infants or older individuals with skin frailty.

## Methods

### Fabrication and characterization of sampling microneedles (MNs)

PDMS molds (Sylgard 184, Dow-Corning) for fabrication of MN patches were prepared by laser micromachining (Blueacre Technology, Ireland). Poly(L-lactide) (PLLA; RESOMER L 207 S, Evonik Industries AG) was melted over the molds under vacuum (−20 mmHg, 200°C, 120 min). Patches were treated with oxygen plasma (Diener Electronic, Germany, 2 min under 0.5 mbar pressure) and then an 0.58% w/v aqueous solution of alginate (PRONOVA SLG20, SLM20, SLG100 or SLM100, Novamatrix, IFF) containing 4.6% v/v sucrose (Teknova, S00572PK) was pipetted onto each patch and allowed to dry at 25°C for at least 4 hours. A cross-linking solution containing 20 mM CaCl_2_ (Sigma, 21115) was then pipetted onto the surface of MN patches and dried in a tissue culture hood overnight.

Microneedle patches were characterized by scanning electron microscopy (SEM) using a Zeiss Crossbeam 540 SEM/FIB. For samples applied *in vivo*, SEM was performed after fixing the samples with paraformaldehyde (4%; Electron Microscopy Sciences, 157-4) and glutaraldehyde (2.5%; Sigma, G7776) for 2 hours and incubated in osmium tetroxide (0.5%; Electron Microscopy Sciences, 19152) for 1 hour before serial dehydration in ethanol. Samples were then dried overnight and imaged. The *in vitro* absorption capacity of the patches was assessed by immersing them in PBS and placing them in a 37°C incubator for a duration ranging from 30 minutes to 18 hours, followed by the measurement of the absorbed liquid mass. For *in vivo* swelling measurements, the patches were weighed both before and after 18 hr insertion into the skin of mice.

### Mice

Animal studies were approved by the Massachusetts Institute of Technology (MIT) Institutional Animal Care and Use Committee, and animals were cared for in the U.S. Department of Agriculture– inspected MIT Animal Facility under federal, state, local, and National Institutes of Health guidelines for animal care. Female, 6-8 weeks C57BL/6 mice (B6(Cg)-Tyrc^-2J^/J, Jax 000058) and KikGR transgenic mice, Tg(CAG-KikGR)33Hadj/J (JAX 013753)^77^, were obtained from the Jackson Laboratory, and colonies were maintained at the animal Koch Institute mouse facility at MIT. Photoconversion was performed as previously described^37^. The skin was exposed to 405-nm LED light from a fixed distance of 1 cm for 3 minutes at 50% intensity, with 5-second breaks every 20 seconds. Black cardboard and aluminum foil were used to shield the rest of the mouse during exposure.

### *In vitro* T cell mobility measurements

Activated T cells were introduced to various alginate groups and then placed inside a Chambered Coverglass (ThermoFisher, 155360). Wells were incubated in Live Cell Imaging Solution (Invitrogen, A59688DJ) and images were acquired using a Leica SP8 laser-scanning confocal microscope equipped with a 25X water lens. 5 FOV were imaged for 35 minutes for each condition under identical settings and subsequently processed using Imaris v10 image analysis software. T cell movement was tracked using the Spots algorithm within the software. At least 50 tracks per condition were analyzed.

### Skin application of sampling microneedles

Animals were anesthetized using isofluorane, and MNs were placed on their back after shaving with an approved depilatory cream. Sampling MNs were laid out flat on 3M Nexcare waterproof tape and subsequently applied by pressing down vertically with the thumb or index finger while securing Nexcare tape around the MN to keep it securely in place. Another layer of waterproof tape was secured around the first layer to keep the MN application site dry during the application period.

### Model protein immunizations

For model protein immunizations, 6-8 week old mice were immunized via subcutaneous injection (at the tail base) of Ovalbumin (OVA, 2.5-10 μg, Worthington) along with an Lipid-conjugated CpG^78^ (Lipo-CpG, 0.4-1.24 nmol, Oligo Factory,), a TLR9 agonist, as an adjuvant. To induce a classic delayed-type hypersensitivity reaction or establish T_rm_ populations on the skin, OVA-immunized mice were intradermally injected with 10 μL of OVA and adjuvant in the flank. For T_rm_ cell recall, mice were intradermally injected with 10 μL of OVA and adjuvant at the same location where T_rm_ had been initially established. To block the infiltration of T cells, mice were treated intravenously with anti–LFA-1 (200 μg per mice, clone M17/4, eBioscience, 14-0111-82). Microneedle sampling, blood draws and skin biopsies were obtained at different intervals. For all in vivo studies, mice were randomly assigned to groups, each consisting of four or more individuals per treatment.

### mRNA synthesis

A set of T cell Simian Immunodeficiency Virus sequences containing putative T cell epitopes^79^ (**Supplementary Table S1**) each separated by an AAY proteasomal cleavage motif were cloned into a DNA plasmid backbone using In-fusion cloning. The plasmid sequence was verified using zero-prep sequencing (Primordium Labs, Arcadia, CA). The resultant plasmid DNA was linearized via HindIII-HF (New England Biolabs, Ipswich, MA) endonuclease digestion and purified with PureLink PCR Purification columns (ThermoFisher, Waltham, MA) following the manufacturer’s instructions. To synthesize mRNA, 20μL *in vitro* transcription (IVT) reactions were performed using the HiScribe T7 High Yield RNA Synthesis Kit (NEB #E2040S), Cleancap AG (TriLink BioTechnlogies, San Diego, CA) and 1–2 μg of linear DNA template. N1-methylpseudouridine (TriLink BioTechnologies, San Diego, CA) was used for the IVT reaction in place of uridine. The IVT product was purified using PureLink RNA Mini columns (ThermoFisher, Waltham, MA) following manufacturer’s instructions. The quality of the resulting mRNA was assessed using UV-Vis spectrophotometry and gel electrophoresis.

### LNP formulations

Lipid nanoparticles (LNPs) were formulated using flash nanoprecipitation in a microfluidic mixer. Briefly, SM-102 ionizable lipid (BroadPharm, San Diego, CA), cholesterol (Avanti Polar Lipids, Snaith, UK), DSPC (Avanti Polar Lipids, Snaith, UK), and PEG-DMG (Avanti Polar Lipids, Snaith, UK) were dissolved in ethanol at a molar ratio of 50:38.5:10:1.5, respectively. mRNA was dissolved in 10 mM citrate buffer pH 3.0 (Alfa Aesar, Haverhill, MA). The lipid and RNA solutions were mixed together at N/P ratio of 6:1 and final RNA concentration of 0.1 mg/mL using an Ignite NanoAssemblr (Precision NanoSystems, Vancouver, Canada) at a flow rate of 12 mL/min and a lipid:RNA flow ratio of 3:1. Formulated LNPs were dialyzed in PBS for 2 hr in 3500 MWCO Slide-A-Lyzer dialysis cassettes (Thermo Fisher, Waltham, MA).

### mRNA Immunization

Mice were injected intramuscularly with mRNA-LNPs encoding T cell SIV epitopes. Three weeks after injection, spleens were excised and splenocytes were isolated using mechanical dissociation. ELISPOT was conducted on the splenocytes using mouse IFNγ ELISPOT kit as per the manufacturer’s protocol. Briefly, well plates were coated with mouse IFNγ and 106 splenocytes/well were seeded on coated wells. Cells were stimulated with SIV peptides at a dose of 2 μg/mL and incubated overnight at 37 °C. Plates were developed according to manufacturer’s protocol and imaged using the CTL-ImmunoSpot Plate Reader. Number of spots were calculated using CTL ImmunoSpot Software. To establish and recall T_rm_ cells, mice received intradermal injections on days 21 and 28, respectively, with subsequent collection of skin and microneedle samples for further analyses.

### Cytokine analysis and flow cytometry

For cytokine sampling, MN patches were placed on the skin for 20 minutes, and for cell sampling, MN patches were placed for 6, 18, or 24 hours. After retrieving the MN patches from skin, they were immersed in 200 ul of PBS containing 2% bovine serum albumin and 100 mM EDTA and incubated at room temperature on a shaker at 150 rpm for 20 min. The supernatant was collected and centrifuged to pellet cells. For cytokine analysis, MN patches were washed in the absence of FBS. The levels of interstitial fluid cytokines and chemokines were determined by a bead-based multiplex assay using the LEGENDplex mouse cytokine response panel kit (740622, BioLegend, USA), following the manufacturer’s protocol, and samples were subsequently subjected to flow cytometry analysis on either an LSR Fortessa or Symphony (BD Biosciences). The concentration of the different cytokines was calculated using the software LEGENDplex V8.0 supplied by BioLegend. For surface staining, cells were incubated with anti-Fc receptor antibody (clone 2.4G2) and stained with antibodies in PBS and 2% fetal calf serum for 30 min on ice. All flow cytometry was conducted on either an LSR Fortessa or Symphony (BD Biosciences), data were collected in Diva (BD Biosciences) and analyzed using FlowJo (BD Biosciences). Antibodies used in these studies: CD45 (BUV395, Cat# 564279, Lot# 2259805, BD Biosciences), CD3L (AF488, Cat# 100321, Lot# B366226, BioLegend), CD3L (PerCP Cy5.5, Cat# 100218, Lot# B365335, Biolegend), CD4 (BUV496, Cat# 612952, Lot# 1328397, BD Biosciences), CD8 (BV421, Cat# 100738, Lot# B358297, BioLegend), CD11b (BV786, Cat# 101243, Lot# B361001, Biolegend), CD103 (BUV 805, Cat# 741948, Lot# 3033478, BD-Optibuild), CD69 (PE cy7, 104512, B320104/B253212, Biolegend), and SIINFEKL/H-2Kb peptide–MHC (major histocompatibility complex) tetramer [iTAg Tetramer/PE–H-2Kb OVA (SIINFEKL) from MBL. Viability was assessed by staining with fixable Live/Dead Zombie NIR (BioLegend).

### Human study design

A healthy individual was recruited at the University of Massachusetts Chan Medical School under Institutional Review Board-approved protocols (STUDY00000321 and H00021295). The volunteer underwent sensitization with squaric acid dibutyl ester (SADBE) in acetone base (Boulevard Compounding Pharmacy, Worcester, MA). In brief, allergic skin reactions were induced by occluding SADBE on the skin using Finn Chamber AQUA patch test chambers (SmartPractice Canada, Calgary, AB, Canada). The following sites were induced and samples were collected at specific timepoints: 1) a site naive to SADBE, sampled two days later; 2) a site previously exposed to SADBE, sampled two days later; 3) a site naive to SADBE, sampled four days later; and 4) a site previously exposed to SADBE, sampled four days later. Additionally, a non-SADBE exposed site was selected as 5) nonlesional skin for comparison.

### Human microneedle sampling

For each site, two sets of MN were firmly secured to the skin using hypoallergenic Scanpor tape (SmartPractice Canada, Calgary, AB, Canada). The first set of MN patch was applied overnight for cell sampling. After removal from the skin, these patches were immersed in 500 μL of PBS with 100 mM EDTA and 2% fetal bovine serum, and then subjected to agitation on a plate shaker for 20 min at 350 rpm. The fluid was subsequently centrifuged at 4°C at 300g for 10 min to pellet the cells. After discarding the supernatant, the pelleted cells were processed for flow cytometry analysis. The second set of MN patch was applied for 20 min to collect extracellular fluid. After removal from the skin, these patches were placed in 200 μL of PBS with 100 mM EDTA and subjected to agitation on a plate shaker for 20 min at 350 rpm. The fluid was then frozen at −20°C and stored for subsequent proteomic analysis.

### Human suction blister skin sampling

Suction blisters were induced on each site using the Negative Pressure Instrument Model NP-4 (Electronic Diversities, Finksburg, MD), with a negative pressure set between 178-255 mmHg until blisters formed. Once formed, blister fluid was aspirated using a 1.0-ml insulin syringe, carefully transferred to a collection tube, and then centrifuged at 4°C at 300g for 10 min to pellet the cells. The supernatant of the blister fluid was subsequently frozen at −20°C and stored for further proteomic analysis. Pelleted cells from the blister fluid were processed for flow cytometry analysis.

### Proteomics analysis

Suction blister and MN patch fluids were analyzed using the Olink Flex platform (Uppsala, Sweden) with a custom panel of targets (see **Supplementary Table S2**). The resulting Olink Normalized Protein Expression (NPX) were converted to pg/mL values and plotted using Prism.

### Statistical analysis

Datasets were analyzed the unpaired Student’s t-test or one-way ANOVA tests, followed by Tukey’s HSD test for multiple comparisons with Prism (GraphPad Software). P values less than 0.05 were considered statistically significant. All values are reported as means ± SEM.

## Data availability

The main data supporting the results in this study are available within the paper and its Supplementary Information. The raw and analyzed datasets generated during the study are available for research purposes from the corresponding authors on reasonable request.

## Acknowledgements

We thank Luciano Santollani for the guidance on the KikGR mice, Coralie Backlund and Alex Hostetler for helping with immunoassays, and Jonathan Dye for assisting with mRNA synthesis. We thank Daniel Garafola for assisting with animal studies and Soo-Yeon Kang for helping with photography. We thank Mariane Melo for her valuable advice on *in vitro* and *in vivo* assays. We thank the Koch Institute Swanson Biotechnology Center for technical support, specifically the Flow cytometry and microscopy core facilities. This work was supported by the NIH (award U01AI176310 to J.H., D.J.I., and S.J.), the Jackson Laboratory, the Ragon Institute of MGH, MIT and Harvard, and by the Koch Institute Support (core) Grant P30-CA14051from the National Cancer Institute. We thank the King Trust, Bank of America Private Bank, Co-Trustees Fellowship, for supporting K.A. D.J.I. is an investigator at the Howard Hughes Medical Institute. All schematic figures were created with BioRender.com.

## Author contributions

S.J., P.T.H. and D.J.I. conceived and designed the experiments. S.J., and R.R.H. conceived the *in vitro* and *in vivo* experiments. S.J., W.K., K.A., N.H., A.K.D., M.R., and J.H. conceived the human microneedle sampling studies. W.K. and K.A. assisted with the human suction blister skin sampling S.J., W.K. and K.A. analyzed the human patient’s data. N.C. aided with LNP formulations and mRNA Immunization. L.M. assisted with the *in vitro* T cell mobility measurements. G.D.G. provided SIV epitopes for generating the mRNA-LNPs. S.J. and D.J.I. wrote the manuscript with contribution from W.K. and K.A. for the human sampling experiments. All authors reviewed and edited the manuscript. S.J., P.T.H., J.H. and D.J.I. supervised the studies.

## Competing interests

S.J., P.T.H. and D.J.I. have submitted a patent application filed by MIT related to the data presented in this work. M.R. is principal or co-investigator of studies sponsored by Pfizer, Biogen, AbbVie, Incyte, LEO Pharma, Abeona Therapeutics, Dermavant, and Target RWE; and M.R. provides consulting for Pfizer, Biogen, Incyte, Takeda, Inzen, ROME Therapeutics, Almirall, Medicxi, Related Sciences, and VisualDx. The other authors declare no interests.

## Notes

### Author Declarations

This study was approved under the University of Massachusetts Chan Medical School Institutional Review Board-approved protocols STUDY00000321 and H00021295

