## Supplementary Information for "Leveraging tissue-resident memory T cells for non-invasive immune monitoring via microneedle skin patches"

### Table of contents

1. Supplementary Tables

2. Supplementary Figures

### Supplementary Tables

#### Supplementary Table S1 | SIV epitopes included in mRNA vaccine

| Region | Sequence |
|---|---|
| **Gag 173-191** | GIRYPKTFGWLWKLVPVNV |
| **Nef 164-182** | YVHLPLSPRTLNAWVKLIE |
| **Gag 144-162** | PDWQDYTSGPGIRYPKTFG |
| **Nef 154-172** | IRFRYCAPPGYALLRCNDT |
| **Env 230-248** | CAPPGYALL |
| **TdaP helper epitope** | GPGPGILMQYIKANSKFIGIPMGLPQSIALSSLMVAQ |

**Supplementary Table S2 | Olink Flex custom panel of targets**

| Assay | Uniprot ID | OlinkID |
|---|---|---|
| **IFNB1** | P01574 | OID15109 |
| **CXCL9** | Q07325 | OID15433 |
| **IFNA2** | P01563 | OID15106 |
| **CXCL13** | O43927 | OID15046 |
| **IL1A** | P01583 | OID15115 |
| **TGFB1** | P01137 | OID15091 |
| **IL33** | O95760 | OID15073 |
| **IL17A** | Q16552 | OID15475 |
| **IL13** | P35225 | OID15310 |
| **TNFSF10** | P50591 | OID15367 |
| **CXCL10** | P02778 | OID15130 |
| **IFNG** | P01579 | OID15112 |
| **CCL19** | Q99731 | OID15556 |
| **TNF** | P01375 | OID15103 |
| **IL15** | P40933 | OID15334 |
| **CXCL8** | P10145 | OID15193 |
| **IFNL1** | Q8IU54 | OID15502 |
| **TNFRSF9** | Q07011 | OID15430 |
| **CXCL11** | O14625 | OID15013 |
| **CCL20** | P78556 | OID15394 |
| **IL4** | P05112 | OID15139 |

### Supplementary Figures

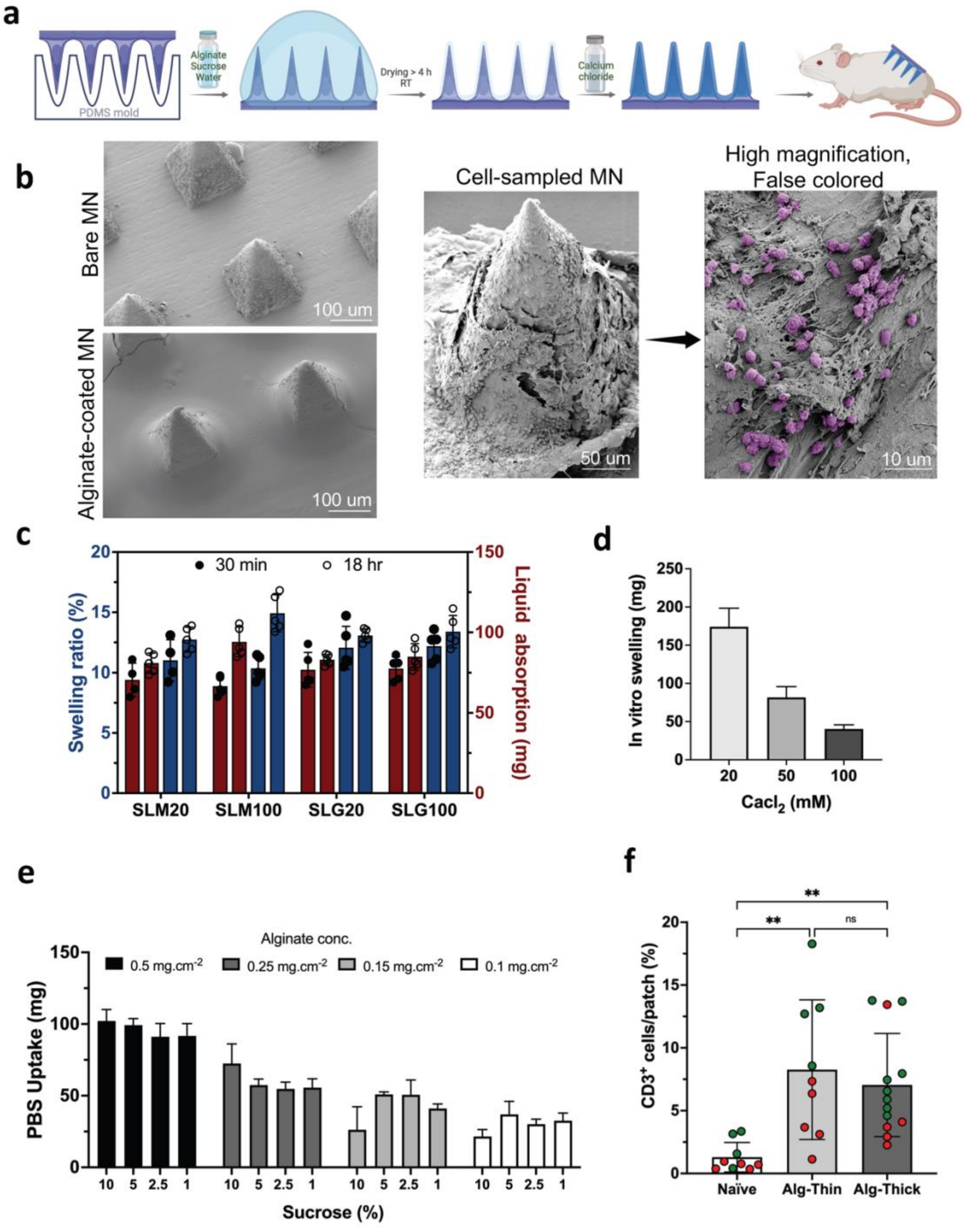


**Supplementary Figure 1**. **a**, Schematic representation of the fabrication and alginate coating process of the microneedle patches. **b**, Scanning electron micrographs of uncoated (bare) and alginate coated MN patches. Electron micrographs of collected cells in SLG20 patch after insertion into the mouse skin. Cells were false-colored in purple to distinguish from the MN base and hydrogel. **c**, In vitro swelling properties of MN patches coated with different alginates. **d, e**, In vitro PBS uptake by SLG20 MN patches as a function of crosslinking concentrations, alginate concentrations, and sucrose content. f, Enumeration of CD3⁺ T cells collected in thin and thick SLG20 MN patches. Green and red dots represent two separate experiments.

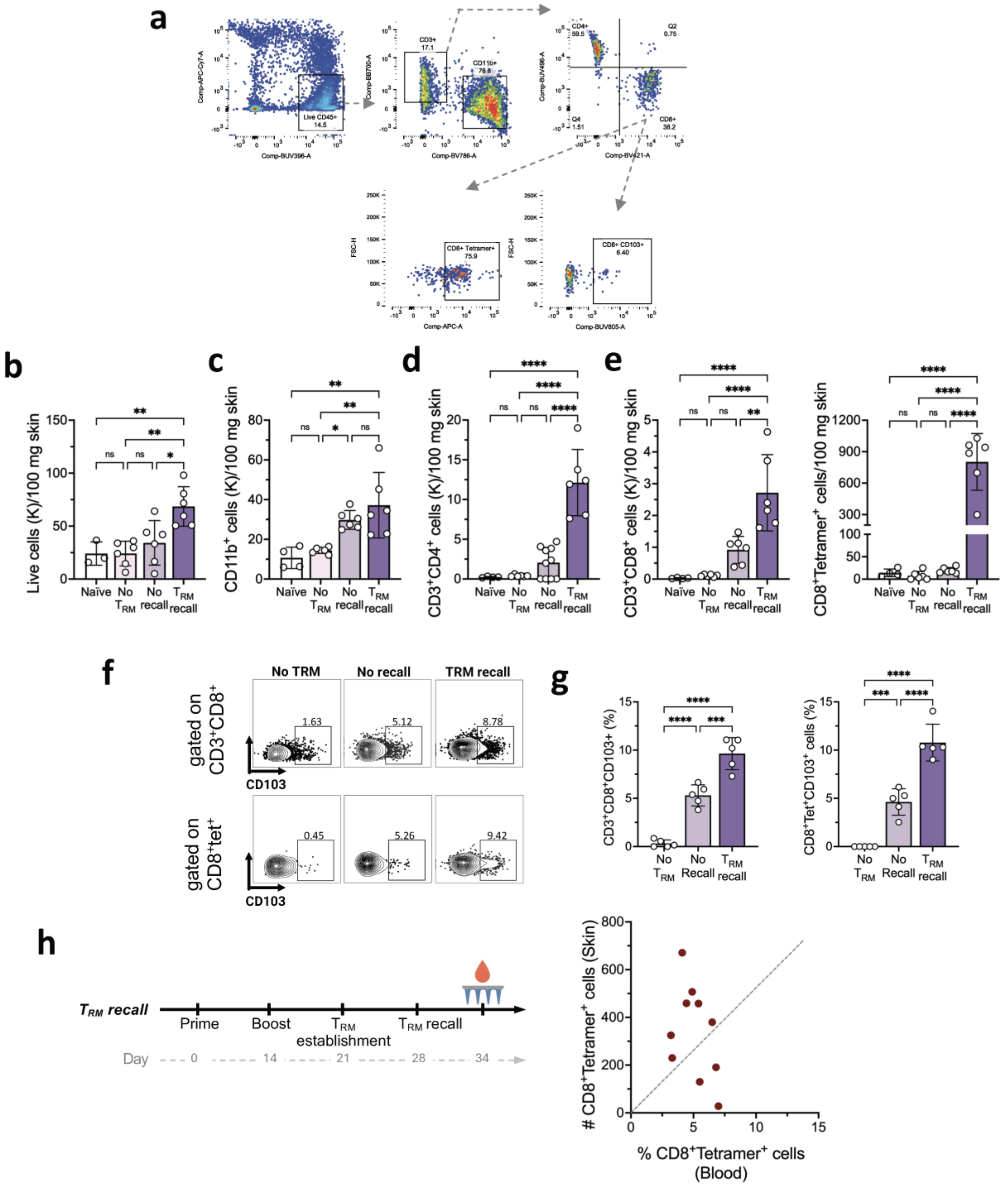


**Supplementary Figure 2**. **a**, representative flow cytometry plots for identifying and quantifying immune cells in the MN patch. **b-e**, Enumeration of recovered total live leukocytes, myeloid cells, T cells, and antigen-specific T cells sampled using skin biopsies. **f,** Representative flow cytometry plots showing expression of CD3⁺CD103⁺ T cells and antigen-specific CD8⁺CD103⁺ $T_{RM}$ cells in the skin. **h**, Enumeration of recovered CD3⁺CD103⁺ T cells and antigen-specific CD8⁺CD103⁺ $T_{RM}$ cells in the skin at $T_{RM}$ establishment and recall steps and in comparison with No $T_{RM}$.

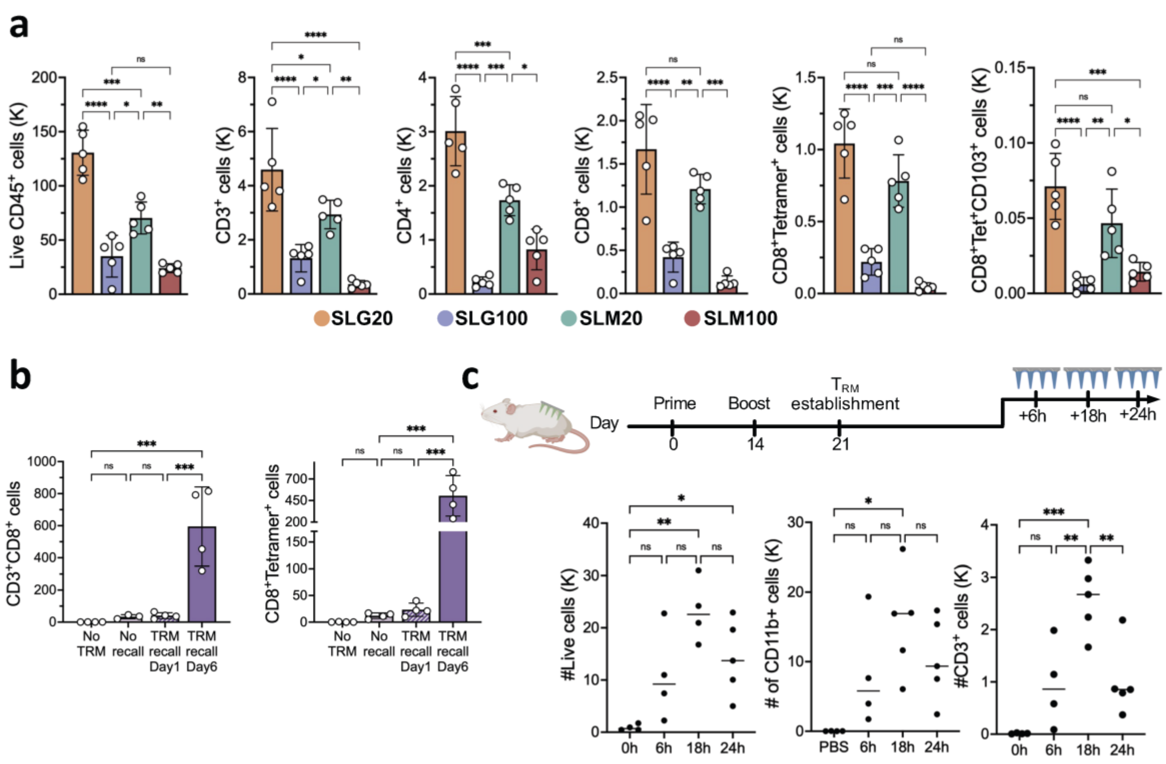


**Supplementary Figure 3**. **a**, Enumeration of recovered total live leukocytes, myeloid cells, T cells, $T_{RM}$ cells, and antigen-specific $T_{RM}$ cells in MN patches coated with different alginates. **b**, Temporal comparison of CD3⁺CD8⁺ T cells and antigen-specific T cells collection 1 day and 6 days post-TRM recall. **c**, Effect of microneedle (MN) patch insertion duration on the number of live cells, myeloid cells, and T cells collected by MN patches.

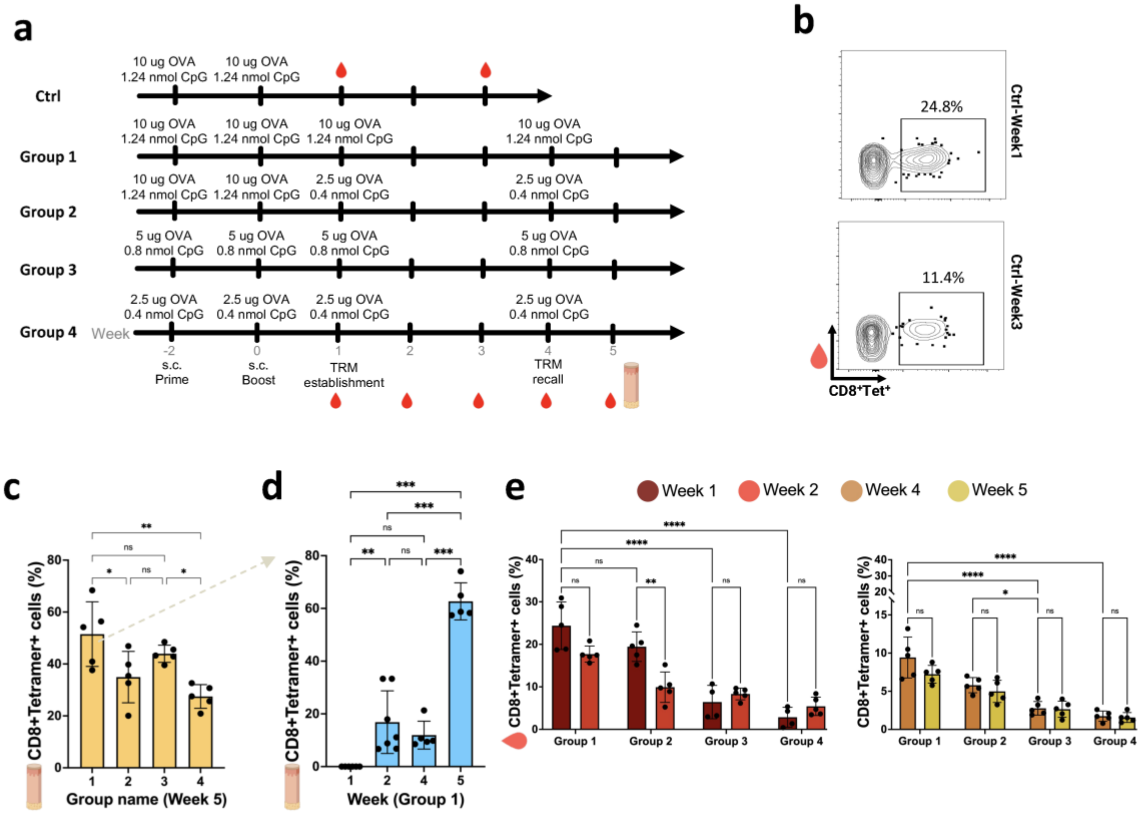


**Supplementary Figure 4**. **a**, study design to compare the effect of multiple amounts of OVA and adjuvant during the prime/boost and $T_{RM}$ establishment/recall steps on the number of antigen-specific lymphocytes systemically and locally. **b**, Representative flow cytometry plots of antigen-specific $CD8^+$ T cells in the control group at 1 and 3 weeks post-prime/boost vaccination. **c, d**, Quantification of antigen-specific T cells in the skin at the injection site across different vaccination regimens and at various intervals after $T_{RM}$ establishment dose. **e**, Quantification of antigen-specific T cells in blood following different doses of $T_{RM}$ establishment and after $T_{RM}$ recall dose.

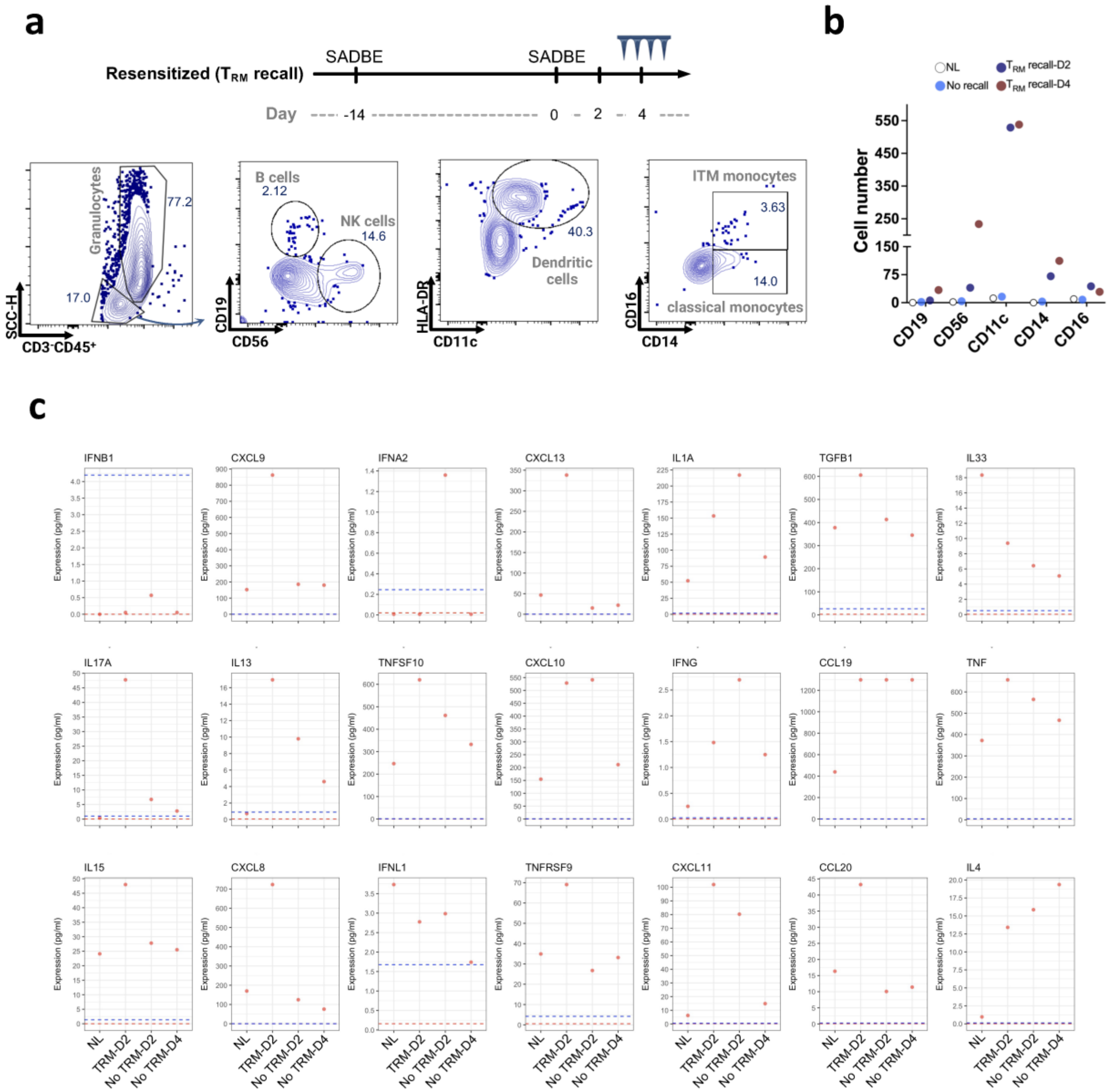


**Supplementary Figure 5**. Olink proteomics data showing temporal changes in skin-associated proteins collected via suction blister method. Key proteins related to humoral immunity are highlighted.
